# Feasibility of an Innovative Absorbable Ventilation Tube Designed to Provide Intermediate-Term Middle Ear Ventilation

**DOI:** 10.1101/2021.05.25.21254504

**Authors:** Sandra Skovlund, Shelagh Cofer, Heather Weinreich

**Affiliations:** University of Minnesota Department of Otolaryngology- Head and Neck Surgery, Minneapolis, MN; Chair- Division of Pediatric Otolaryngology, Mayo Clinic, Rochester, MN; University of Chicago

**Keywords:** feasibility, eustachian tube dysfunction, ventilation tube, absorbable, middle ear ventilation

## Abstract

Myringotomy with ventilation tube placement is a common surgical procedure performed in children and adults to remove fluid build-up behind the tympanic membrane. However, retention of tubes beyond achievement of therapeutic response increases risk for complications and additional intervention. This small feasibility study was conducted to demonstrate proof-of-concept of a novel bioabsorbable ventilation tube that provides the necessary duration of ventilation with absorption shortly thereafter in humans. Tubes were placed in 15 ears of 14 patients meeting indications for short or intermediate-duration of middle ear ventilation. Two independent examiners documented tube patency and tube absorption status at 3, 6 and 12 weeks or until absorption was complete. Results indicate that average ventilation time was 12 weeks (range 3 weeks to 18 months). There was no observation of blockage. These findings support the feasibility of a novel bioabsorbable ventilation tube.

## INTRODUCTION

Myringotomy and ear tube placement is a common surgical procedure to remove or prevent fluid build-up behind the eardrum or to instill medication into the middle ear. Placement of ear tubes, also referred to as ventilation tubes, is the most common surgery performed in the pediatric population for repeated middle ear infections. Ear tube placement is also indicated in adults with eustachian tube dysfunction, serous otitis media(SOM), sudden sensorineural hearing loss(SSNHL) or a Meniere’s exacerbation(ME). Previous reports suggest that ventilation time of 3 weeks can be sufficient to prevent recurrence of otitis media in a majority of cases.^1^ In most cases the ventilation tube self-extrudes at 6-18 months, however complications have been reported if durable tube material(fluoroplastics, silicone, titanium) remains in place beyond 4-6 months.^2,3^ In practice, tubes that are retained beyond two years are removed and the residual perforation is patched.

To address the need for an ear tube that provides intermediate-term ventilation to successfully treat several ear conditions without risks of foreign body complications, a bioabsorbable ventilation tube (BVT) has been developed. The BVT is constructed from materials commonly used in the ear that have substantial documentation of safety and bioabsorption predictability. The objective of this study was to evaluate feasibility of this novel BVT in adult patients indicated for intermediate-duration middle ear ventilation.

## METHODS

This small feasibility study was conducted at a single multi-specialty group practice was approved by an Institutional Review Board for Human Subjects Research. Adult participants with acute SOM, SSNHL, or ME were eligible for placement of a short-duration BVT tube in a clinic setting. The BVT is composed of gelatin and a suspension of antibiotic and steroid that is commonly administered in the middle ear. Patients were evaluated at weeks 3, 6, and 12 by two examiners to document time to BVT lumen patency in the process of absorption. Exam findings were verified and recorded independently by each examiner using binocular microscopy.

## RESULTS

Fourteen participants, ages 26-80 years (mean age 61 years, 47% female) voluntarily underwent placement of a BVT tube in 15 ears. All patients completed the placement and absorption period without complication. Two patients were not included in the descriptive statistics for ventilation time: 1 patient (1 ear) who did not complete evaluation at the designated intervals; and 1 patient (2 ears) who received radiation following BVT placement and had an expected delay in tube absorption.

Average time for middle ear ventilation was 9 weeks (range 3 weeks to 12 weeks; n=12). BVTs showed progressive signs of resorption at each follow-up visit. There were no observations of BVT lumen occlusion.

## DISCUSSION

This report provides feasibility for a novel bioabsorbable ventilation tube in adults who met criteria for short or intermediate-term ventilation of the middle ear. Ventilation tube placement is a common treatment for numerous ear conditions in pediatric and adult populations. In the interval between tube placement and 4-6 months, ventilation tubes are quite successful at clearing otitis media, yet ventilation tubes made with durable materials(fluoroplastics, silicon, titanium) have been shown to cause expensive complications starting at 4-6 months and following tube removal at 2 years post-placement due to a higher rate of eardrum perforations.^2^ These complications contribute to additional health care costs ($170 -$3000 per case) related to otorrhea, ear drum perforation, tube obstruction, granulation tissue/foreign body reaction, and segmental atrophy.^2,3^ The adequate duration of ventilation to re-establish normal middle ear function has yet to be established, though there is evidence time to efficacy is a shorter time than duration of current durable tubes treatment. Studies have shown that ventilation of less than 4 months is adequate for treatment of recurrent otitis,^1^ indicating the opportunity to reduce over-treating without compromise to the primary clinical outcome and with a potential benefit of reducing complications.

The BVT development and application presented here is aligned with medical advances focusing on minimally invasive interventions and efficient clinic utilization, including the exploration of absorbable devices to treat disease. In endoscopic sinus surgeries, drug-eluting absorbable stents have been successful in decreasing scarring and polyp recurrence in the postoperative period following sinus surgery.^4^ Furthermore, absorbable facial plating for pediatric trauma is used to eliminate a life-long foreign body. Here the BVT is composed of materials already accepted and used by Otolaryngologists, thereby providing the surgeon predictable behavior and reassurance. Our results confirm previous reports of gelatin resorption in the ear without complication.^5^

## CONCLUSION

Here we provide feasibility of a novel BVT that provides adequate ventilation time for potential clinical application in adult patients with assorted otological conditions. In addition to providing potential benefit for traditional indications such as otitis media, the BVT may be indicated in the growing use of eustachian tube balloon dilation, hyperbaric oxygen treatments, and drug delivery to the middle ear. A clinical study will be initiated to optimize BVT utility for these conditions.

## Data Availability

Data regarding device status and functionality is available as documented.

## ACKNOWLEDGEMENTS

The authors thank Amy Moore, PhD for her review of the manuscript.

